# Repeatability and reproducibility of spectral results with clinical photon-counting CT: multi-center phantom study

**DOI:** 10.64898/2026.10.30.26366926

**Authors:** Leening P. Liu, Martin V. Rybertt, Peter B. Noël, Liqiang Ren, Xinhui Duan, Kishore Rajendran, Di Zhang, Grace H. Kim, Michael F. McNitt-Gray

**Affiliations:** Department of Radiology, University of Pennsylvania, Philadelphia, PA, USA; Department of Bioengineering, University of Pennsylvania, Philadelphia, PA, USA; Deparment of Radiology, University of Texas Southwestern Medical Center, Dallas, TX, USA; Department of Radiology, Mayo Clinic, Rochester, MN, USA; Department of Radiological Sciences, University of California, Los Angeles, Los Angeles, CA, USA

## Abstract

**Background:** Photon-counting CT utilizes a new detector technology that enhances quantitative imaging but introduces new system sensitivities that may affect quantitative reproducibility. For biomarkers, this quantitative reproducibility across protocol parameters, between institutions, and over time boosts their generalizability and clinical adoption for improved disease stratification. While stability across protocol parameters and over time have previously been evaluated, inter-institution reproducibility with PCCT remains unknown.

**Purpose:** This study aimed to evaluate the repeatability and reproducibility of PCCT spectral results across four institutions using the same scanner model, phantom model, and protocol parameters.

**Methods:** Four institutions scanned the same material inserts from the same multi-energy CT phantom model on the same PCCT model at a tube voltage of 140 kV and a volumetric dose index (CTDI_vol_) of 10 mGy. For all scans, virtual monoenergetic images (VMI) from 40 to 190 keV, iodine maps, and virtual non-contrast maps (VNC) were analyzed to determine the mean error relative to the expected value for each insert, spectral result, phantom copy, and institution. For all spectral results, the standard deviation across phantom copies and institutions (σ_ip_) was calculated for each material insert to highlight variation from both phantom copies and institutions. Additionally, repeatability (RC) and reproducibility coefficients (RDC) were determined.

**Results:** As VMI energy increased, quantitative variation between institutions decreased and stabilized. At VMI 70 keV, the σ_ip_ was 1.3, 3.8, and 2.7 HU for the blood 70, adipose, and iodine 5 mg/mL, respectively. The σ_ip_ increased to 7.4 HU at VMI 50 keV and decreased to 1.5 HU at VMI 150 keV. Similarly, the RDC for the iodine 5 mg/mL insert decreased from 20.6 to 4.1 HU as VMI energy increased from 50 to 150 keV. Iodine maps and VNC also exhibited reproducibility across the four institutions with RDC ranging from 0.08 to 0.49 mg/mL and 3.5 to 10.9 HU, respectively, across inserts.

**Conclusion:** This multi-institutional study evaluated the variation of PCCT across a range of inserts and spectral results and demonstrated decreasing variation with increasing VMI energy levels. The characterized repeatability and reproducibility not only further reinforce the benefits of PCCT to support clinical translation and adoption but also provide guidance to ensure generalizable and clinically useful biomarkers for a range of clinical applications.

## Introduction

Quantitative imaging augments qualitative interpretation with objective metrics, or biomarkers, extracted from images in order to more finely characterize and stratify disease. The improved stratification of disease enables personalized and targeted clinical management. In CT, biomarkers span morphology^1^, attenuation-based metrics^2–5^, and radiomic features^6–8^ to detect subtle characteristics and changes in biological disease. Quantitative imaging has been promising for a range of clinical applications, from cancer staging and treatment response^9–11^ to coronary plaque characterization^12,13^. While clinical translation has increased over the last decade, clinical adoption has been hindered by modest reproducibility^14,15^. Visualization of key structures and attenuation are affected by key sources of variation, such as patient habitus, scanner model, tube spectra, and protocol parameters^16–18^. This variability impedes quantitative reproducibility across institutions, scanner models, and protocol parameters, thus limiting the generalizability and clinical utility of biomarkers.

Spectral CT has enabled the generation of quantitative maps, or spectral results, that are more agnostic to x-ray beam spectra and reproducible compared to conventional CT images^19^. The enhanced reproducibility stems from the implementation of a two-basis material decomposition. The result is quantitative spectral maps, such as virtual monoenergetic images (VMI), iodine maps, and virtual non-contrast (VNC) maps, that are more robust in differences in the tube spectra and other key protocol parameters, such as radiation dose and body habitus. In particular, conventional CT images represent the attenuation at the effective energy of an unknown and scanner dependent x-ray spectra. VMIs, on the other hand, correspond to the attenuation at a known energy, which can be readily calculated for known material compositions, such as those provided by a phantom with known reference materials. Across spectral results, quantitative stability has been demonstrated across varying tube voltages, body habitus, radiation dose, and iterative denoising levels^20–24^. Additionally, the variability with key protocol parameters has been evaluated on different implementations of spectral CT, including newer generation CTs, such as photon-counting CT (PCCT)^25–27^.

PCCT shifts the paradigm with its unique energy discriminating detector that boosts spatial resolution, enhances quantitative imaging, and facilitates low-dose imaging^26,28–33^. However, the detector may be influenced by ambient temperature and humidity. This could present a new source of variation and/or artifacts that may affect not only previously established benefits of PCCT that support clinical translation but also the quantitative stability and reproducibility required for quantitative imaging. A previous study demonstrated the quantitative stability of spectral results over two years for a range of spectral results, but the inter-institutional reproducibility is still unknown^34^. We aimed to assess the inter-institutional reproducibility of quantitative spectral results across four US-based institutions with the same scanner model, phantom model, and protocol parameters.

## Methods

### Phantom

Each of the four institutions utilized the same model of a commercially available phantom (Multi-energy CT Phantom, Sun Nuclear, Figure 1) described in AAPM TG 299^35^. The phantom has a central cylindrical section with a diameter of 20 cm and an elliptical extension ring with dimensions of 30 x 40 cm. In these two sections, tissue-mimicking and material-specific inserts were placed in the same configuration across the four institutions. The inserts included materials labeled as adipose, brain, blood 40, blood 70, blood + 2 mg/mL iodine, blood + 4 mg/mL iodine, calcium 50 mg/mL, iodine 2 mg/mL, iodine 5 mg/mL, iodine 10 mg/mL, and solid water.

**Figure 1.**
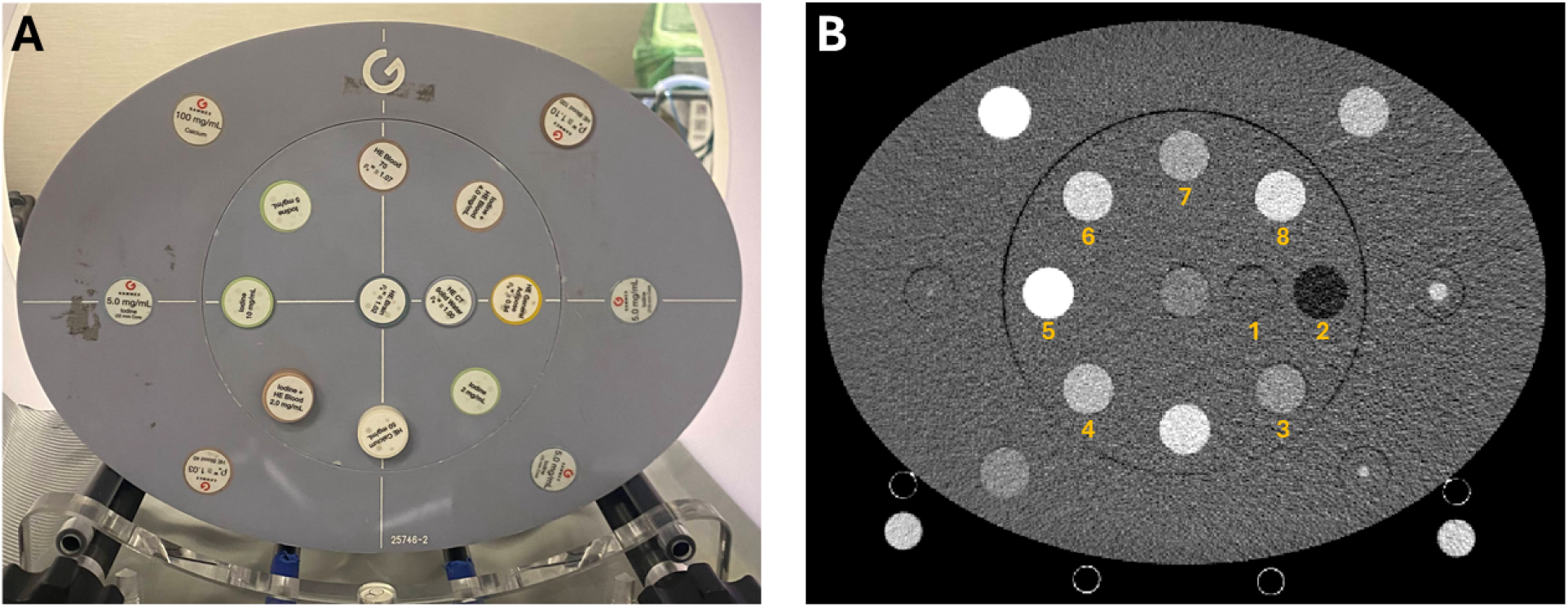
Phantom. A multi-energy CT phantom (A) was utilized to evaluate repeatability and reproducibility of quantitative spectral results across four institutions. The phantom consisted of tissue-mimicking and material-specific inserts for analysis (B): 1. solid water, 2. adipose, 3. iodine 2 mg/mL, 4. blood + 2 mg/mL iodine, 5. iodine 10 mg/mL, 6. iodine 5 mg/mL, 7. blood 70, and 8. blood + 4 mg/mL iodine. WL/WW: 40/300 HU.

To account for potential differences in phantom versions and manufacturing processes, elemental composition and physical density provided by the phantom manufacturer was collected for all four phantoms. Using these specifications and the elemental mass attenuation coefficients from NIST XCOM^36^, expected Hounsfield Units (HU) were calculated for each institution’s insert at energies ranging from 40 to 190 keV to match the available range of VMIs,

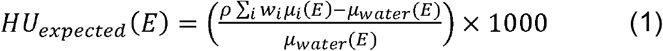

Where *HU_expected_*(*E*) is the expected HU of the insert at energy *E,p* is the physical density of the insert,*w_i_* is the mass fraction of element *i* in the insert,*µ_i_(E)* is the mass attenuation coefficient of element *i* at energy *E*, and *µ_water_(E)* is the mass attenuation coefficient of water at energy *E*. Additionally, the range of expected values for each institution’s inserts was determined by utilizing the manufacturer prescribed physical density tolerance of 0.02 g/mL. For iodine and VNC maps, the expected values of iodine-containing inserts were determined by first calculating the expected attenuation at 60 and 120 keV to provide two sets of attenuations that are well separated in energy. Then, a two-basis material decomposition was performed with water and iodine to extract an expected iodine density and VNC attenuation for each insert. This approach was utilized rather than iodine concentration values provided by the manufacturer to better simulate the realistic processing that is performed to generate iodine maps and VNCs. These expected values for VMIs, iodine maps, and VNCs are later used to calculate the error in measured values.

### Image acquisition and reconstruction

At each institution, the phantom was placed in the isocenter and scanned with a first-generation clinical dual-source PCCT (NAEOTOM Alpha.Peak, Siemens Healthineers; Figure 2, solid lines, “multi-institution”). All scanners were operated with the same software version (VB20) and protocol parameters to eliminate software- and protocol-specific differences. Phantoms were scanned in single-source mode (RoutineSpiralAdultAbdomenQuantumPlus) at a tube voltage of 140 kVp. Scans were repeated three times in order to account for statistical variation. Exposure modulation was turned off; a constant exposure was set for each scan to achieve a volumetric CT dose index of 10 mGy, approximately matching the achievable dose for contrast-enhanced abdomen/pelvis CT^37,38^. For all scans, spectral results, including VMIs from 40 to 190 keV, iodine maps, and VNC, were reconstructed with a field of view of 450 mm, a slice thickness of 2 mm, a reconstruction kernel of Qr48, and denoising (Quantum Iterative Reconstruction, QIR) level of 2 (Table 1).

**Figure 2.**
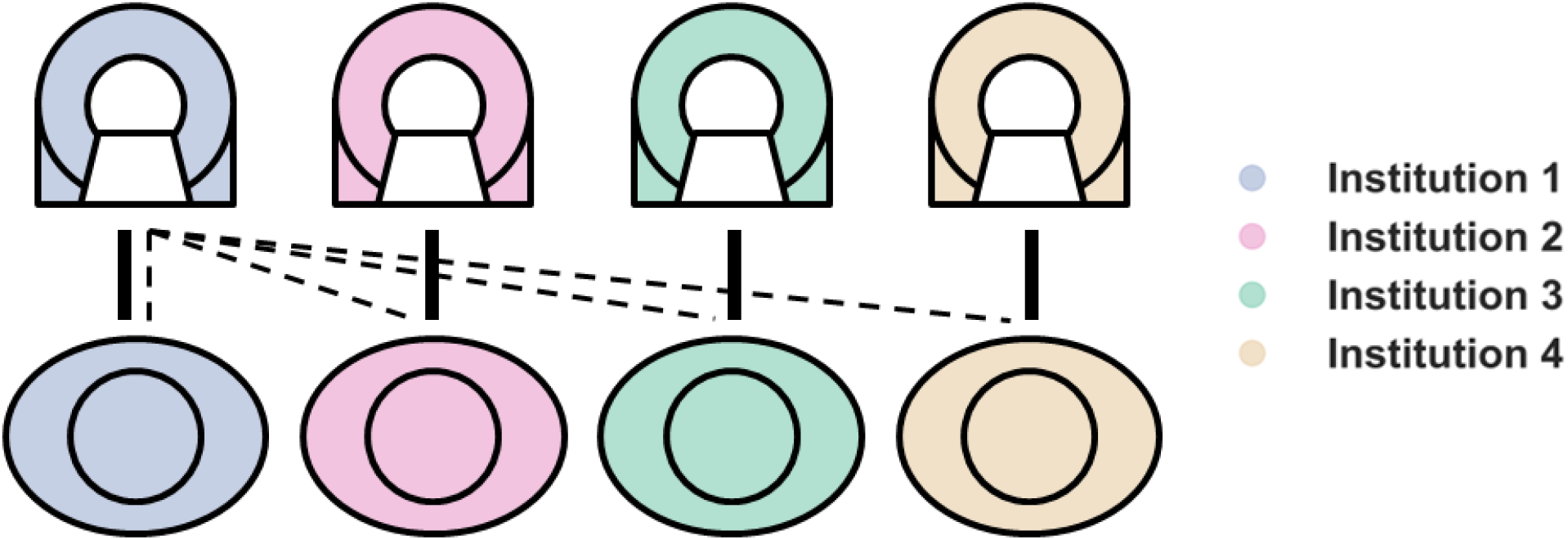
Inter-phantom and institutional evaluation of photon-counting CT. To assess the repeatability and reproducibility of quantitative spectral results from PCCT, four institutions scanned their copy of the same phantom model on the same PCCT model (solid lines). In addition, each phantom’s inserts were independently scanned at a single same institution in order to isolate phantom-specific differences (dashed lines).

**Table 1.**
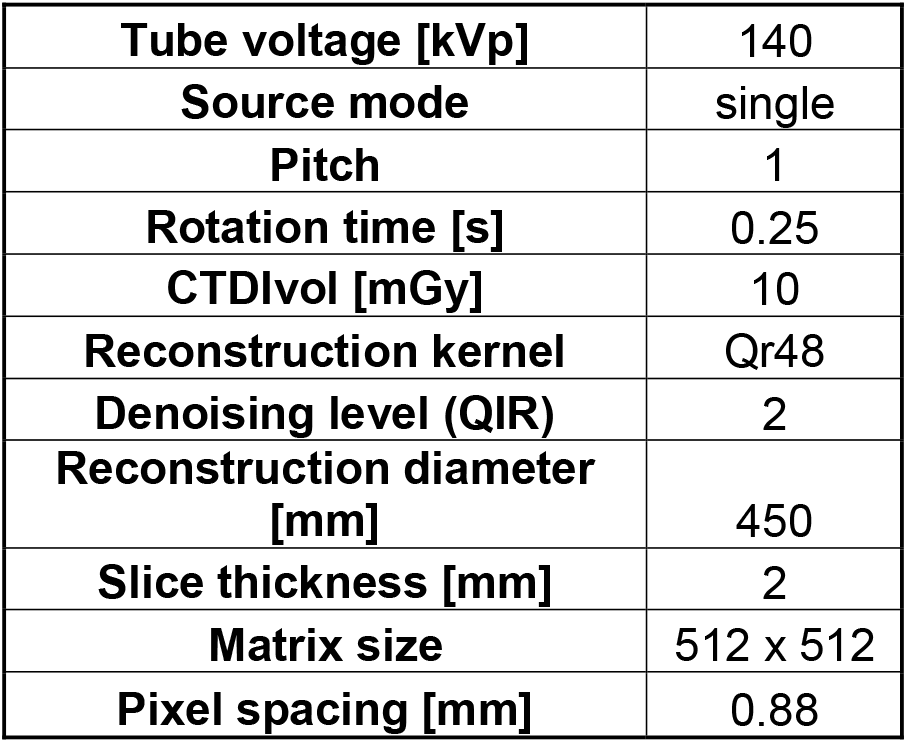
Acquisition and reconstruction parameters.

In addition, phantom-specific variations were evaluated using the PCCT at Institution 1 (Figure 2, dashed lines, “single-institution”). The phantom inserts from each of the participating institutions were scanned within Institution 1’s phantom body using the same phantom configuration to evaluate inter-phantom reproducibility. The same protocol parameters were utilized to isolate phantom-specific variation (Table 1).

### Overall image analysis

For each set of scans that correspond to a phantom and institution combination, in-house software was utilized to determine the center of the inserts on VMI 70 keV images. Using the center of the inserts, regions of interest (ROI) with a diameter of 16.8 mm were placed on all spectral results for the set of scans. The mean value (HU or iodine density) was subsequently measured on the central ten slices for each of the three repeated scans. The error for each phantom’s insert and spectral result (Equation 2) was then calculated as the difference between the measured value and the expected value for the corresponding phantom, insert, and spectral result:

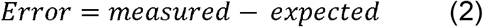

While percent error has been recommended for evaluation of spectral CT systems^35^, the nominal error was utilized to capture variation in HU. The error for each institution, phantom copy, insert, and spectral result was then averaged across image slices and scan repetitions to obtain a mean error. The average error for each insert and institution was represented in a scatter plot across VMI energies. A scatter plot was utilized to visualize the average error for each insert and phantom at the same institution. This visualization was also repeated for average error in iodine maps and VNC. For these scatter plots, error bars corresponded to the standard deviation of error across slices and scan repetitions for each insert, phantom copy, institution, and spectral result.

### Single-institution data analysis

For the single-institution data, the main source of variation is the phantom inserts. To isolate this variation, the following metrics were evaluated:

i. *Variability.* Mean error from the different phantom insert copies was utilized to calculate the inter-phantom standard deviation (σ_p_) for each insert and spectral result.
ii. *Inter-phantom reproducibility.* Inter-phantom reproducibility corresponds to the agreement in error across phantoms at a single institution. The inter-phantom reproducibility coefficient (RDC_p_) was calculated for each insert and spectral result by using a one-way analysis of variance (ANOVA):

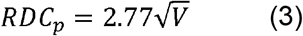

where *V* is the sum of all variance components from the ANOVA except the random effects variance. The RDC_p_ represents the two-tailed 95% confidence interval for repeated measurements between phantoms at a single institution.

### Multi-institution data analysis

For multi-institutional data, the primary sources of variation include both the phantoms and scanners. In addition, variation is present within each scanner. To examine the effect of the different sources of variation, the following metrics were calculated:

i. *Intra-institution and -phantom variability.* The standard deviation across errors for each institution, phantom copy, insert, and spectral result (σ) were determined to characterize the intra-institutional and -phantom variability for each of the four institutions.
ii. *Repeatability.* Repeatability, defined as the agreement in error within the same institution and phantom copy, was quantified using the multi-institution data. σ was utilized to calculate the repeatability coefficient (RC)^39^:

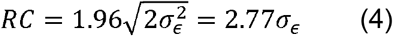

where σ*_ɛ_*^2^ is the variance at a given institution *ɛ*. RC represents the two-tailed 95% confidence interval for measurements repeated at the same institution with their respective phantom for a given institution, insert, and spectral result (intra-institution and -phantom).
iii. *Inter-institution and -phantom variability.* The inter-institution and -phantom standard deviation (σ_ip_) was collectively calculated across the mean error at different institutions for each insert and spectral result. In order to demonstrate the difference in variability from the primary sources of error, the σ_ip_ and σ_p_ were plotted across VMI energies for each insert.
iv. *Inter-institutional and -phantom reproducibility*. Inter-institutional and -phantom reproducibility corresponded to the agreement in error across phantoms and institutions. To calculate reproducibility, a one-way ANOVA was applied to obtain the reproducibility coefficient (RDC_ip_) for every insert and spectral result^39^:

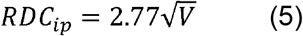

Where *V* is the sum of all variance components from the ANOVA except the random effects variance. The RDC_ip_ represents the two-tailed 95% confidence interval for repeated measurements between institutions and phantoms.

## Results

### Repeatability

Spectral results demonstrated consistent quantification at each institution (Table 2). At VMI 70 keV, σ of the blood 70 insert were 1.0, 1.0, 1.0, and 0.9 HU for the four institutions, respectively. Similarly, the σ of the iodine 5 mg/mL insert at VMI 70 keV were 1.4, 1.2, 1.0, and 0.8 HU for the four institutions, respectively. The corresponding RC for the blood 70 and iodine 5 mg/mL insert at VMI 70 keV ranged from 2.5 to 3.0 and 2.2 to 3.9 HU, respectively (Table 2). As VMI energy decreased to 50 keV, σ of the iodine 5 mg/mL insert increased to 2.2, 1.8, 1.4, and 2.3 HU for the four institutions, respectively. Across inserts, RC also increased: RC ranged from 3.8 to 6.3 HU for VMI 50 keV compared to 2.2 to 3.9 HU for VMI 70 keV (Table 2). At high energy VMIs, RCs were similar to those at VMI 70 keV. For VMI 100 and 150 keV, the minimum RC across inserts was 1.9 and 2.0 HU, while the maximum RC across inserts was 4.5 and 4.8 HU, respectively. With iodine maps, RC ranged from 0.06 to 0.25 mg/mL, demonstrating some variability in repeatability between the institutions. VNC showed similar repeatability to VMI 70 keV with RC ranging from 3.1 to 4.0 HU for the iodine 5 mg/mL insert.

**Table 2.** Range of repeatability coefficient (RC) across institutions for each insert.

| **Insert** | **Range of repeatability coefficient (RC)** |  |  |  |  |  |
|---|---|---|---|---|---|---|
|  | **VMI 50 keV [HU]** | **VMI 70 keV [HU]** | **VMI 100 keV [HU]** | **VMI 150 keV [HU]** | **Iodine [mg/mL]** | **VNC [HU]** |
| Solid Water | [4.6, 5.5] | [2.7, 3.8] | [2.8, 4.5] | [3.0, 4.8] | [0.11, 0.17] | [3.4, 6.6] |
| Adipose | [3.6, 5.5] | [1.9, 3.2] | [2.7, 3.3] | [2.8, 3.7] | [0.08, 0.11] | [3.2, 4.6] |
| Blood 70 | [3.1, 5.5] | [2.5, 3.0] | [1.9, 3.0] | [2.0, 3.1] | [0.06, 0.14] | [2.0, 3.2] |
| Io 2 mg/mL | [4.1, 8.4] | [2.6, 3.2] | [2.7, 4.4] | [2.7, 5.3] | [0.08, 0.25] | [3.0, 7.6] |
| Io 5 mg/mL | [3.8, 6.3] | [2.2, 3.9] | [2.6, 3.5] | [2.7, 3.7] | [0.08, 0.17] | [3.1, 4.0] |
| Io 10 mg/mL | [6.2, 8.4] | [3.1, 4.6] | [3.4, 3.7] | [3.3, 4.4] | [0.14, 0.19] | [3.7, 6.4] |
| Blood + 2 mg/mL Io | [6.1, 7.2] | [3.3, 4.4] | [2.6, 3.5] | [2.6, 3.8] | [0.14, 0.17] | [2.8, 4.0] |
| Blood + 4 mg/mL Io | [5.2, 6.8] | [2.6, 3.0] | [2.7, 3.5] | [2.9, 4.0] | [0.14, 0.17] | [3.1, 4.4] |

### Inter-phantom reproducibility

Spectral results exhibited similar quantification between phantoms scanned at a single institution (Figure 3). With the blood 70 insert, VMI 70 keV errors for each institution’s phantom copy measured 6.7 ± 0.6, 5.0 ± 1.2, 6.6 ± 1.0, and 5.7 ± 1.1 HU (Figure 3). The resulting σ_p_ was 0.8 HU. Across inserts, the σ_p_ for VMI 70 keV ranged from 0.7 to 3.6 HU (Figure 4), corresponding to an average RDC_p_ of 4.3 ± 3.3 HU. Similar to repeatability, variability between the phantom increased with lower energy VMIs. While the error between phantoms remained similar, the σ_p_ of the iodine 5 mg/mL insert increased from 0.9 to 1.8 HU as VMI energy decreased from 70 to 50 keV (Figure 4). On average, the RDC_p_ for VMI 50 keV was 7.1 ± 3.4 HU. Conversely, at high VMI energy, inter-phantom variability stabilized. The σ_p_ across inserts averaged 1.2 ± 1.3 and 1.3 ± 1.2 HU at VMI 100 and 150 keV, respectively (Figure 4). Iodine and VNC also showed phantom variability with RDC_p_ ranging from 0.07 to 0.25 mg/mL and 3.5 to 10.9 HU, respectively.

**Figure 3.**
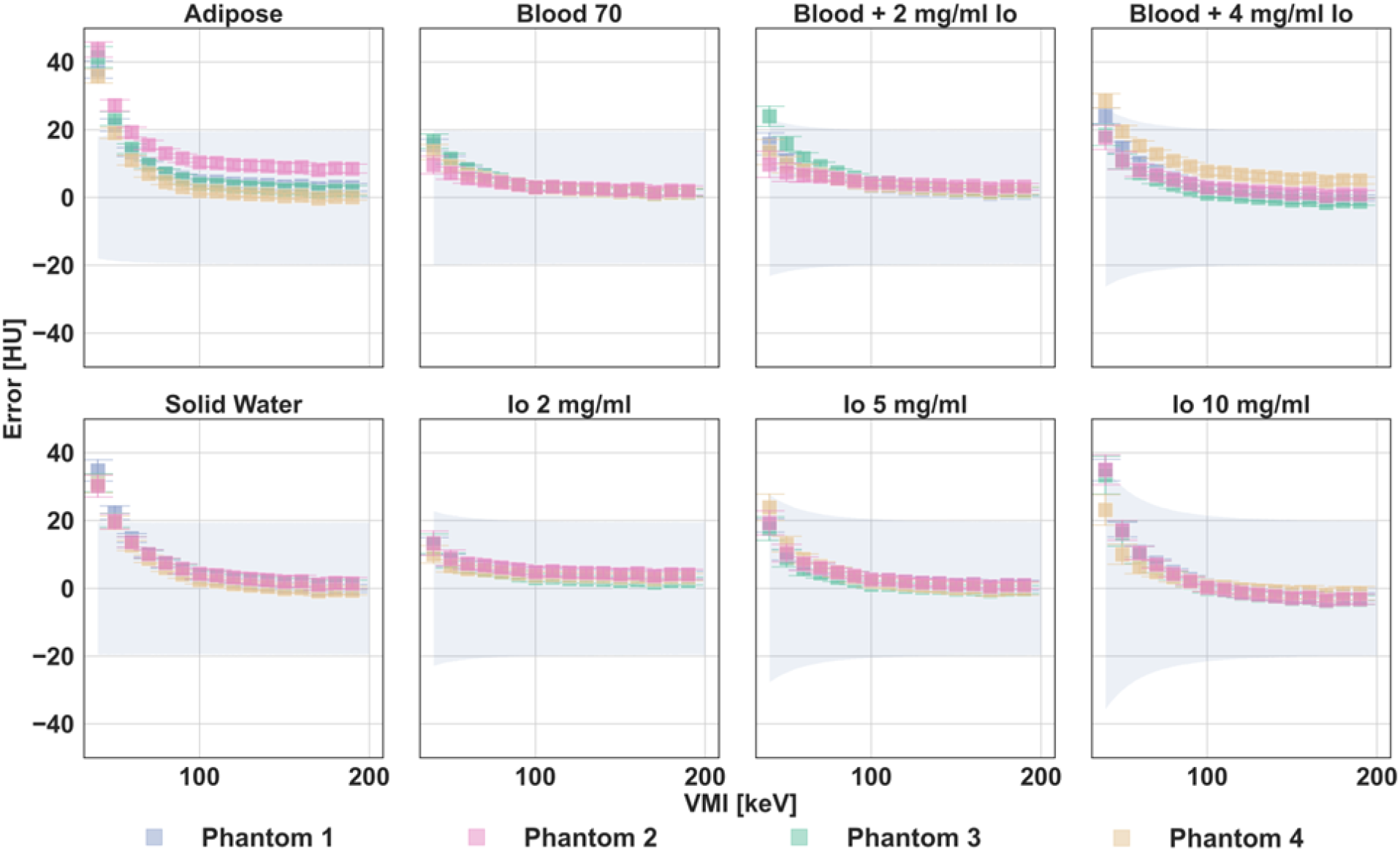
VMI error of different inserts across four phantoms at a single institution. VMI error was similar across the phantoms at a single institution for all insert, indicating minor quantitative differences between phantoms. Error bars represented the standard deviation of the error across slices and scan repetitions with each phantom and institution combination. The blue shaded region corresponded to the expected tolerance of the error as determined by the manufacturer-defined physical density tolerance.

**Figure 4.**
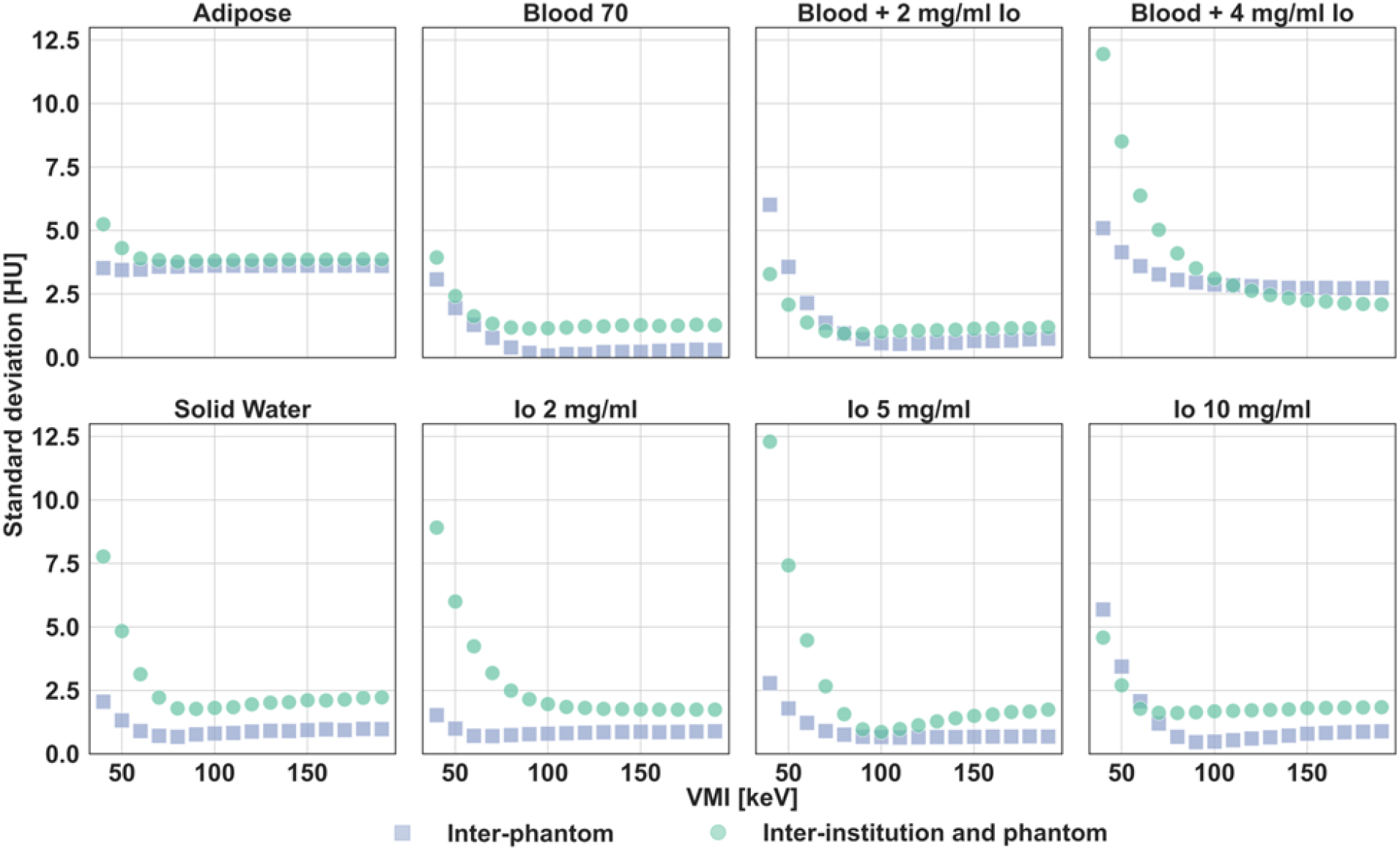
VMI variability between institutions and phantoms. Inter-institution and phantom standard deviation (σ_ip_) were generally larger than inter-phantom standard deviation (σ_p_) across inserts and VMIs. This difference in variability highlights the stronger effect of institution compared to the effect of the phantom.

### Inter-institution and -phantom reproducibility

At VMI 70 keV quantification was consistent between institutions for the different inserts (Figure 5). For the blood 70 insert, errors for each institution and phantom combination were 7.3 ± 1.0, 4.9 ± 1.0, 4.5 ± 1.0, and 6.4 ± 0.9 HU, respectively. The σ_ip_ was 1.3 HU compared to σ_p_ of 0.8 HU, showing a small increase in variability from utilizing each institution’s PCCT (Figure 4). A similar effect was observed in other inserts, including the iodine 5 mg/mL insert where the σ_ip_ and σ_p_ were 2.6 and 0.9 HU, respectively (Figure 4). Across all inserts, the σ_ip_ at VMI 70 keV ranged from 1.0 to 5.0 HU, corresponding to an average RDC_ip_ of 7.2 ± 3.8 HU (Table 3).

**Figure 5.**
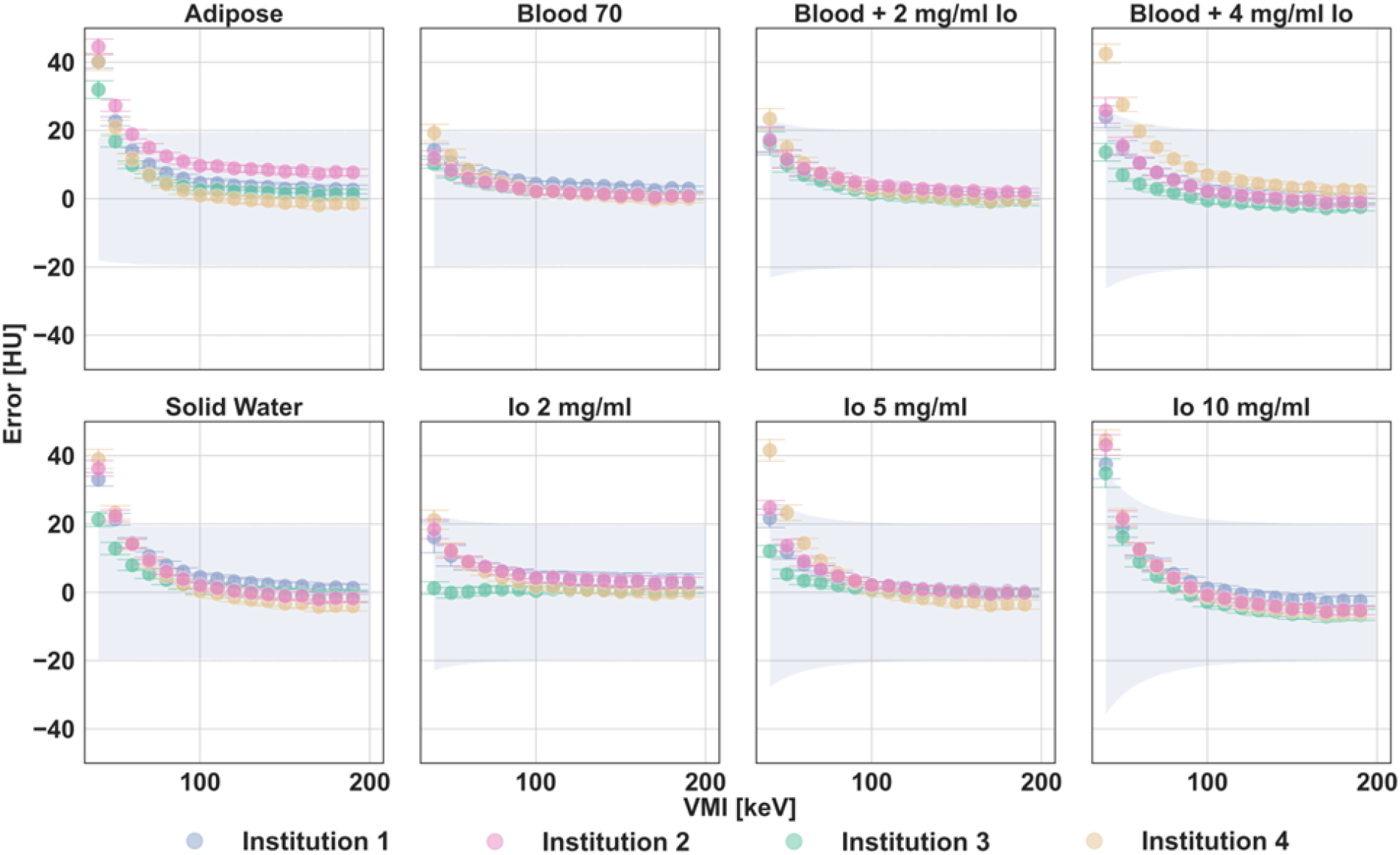
VMI error of different inserts across four institution and phantom pairs. For all inserts, VMI error across institutions became more concordant as VMI energy increased. Error bars represented the standard deviation of the error across slices and scan repetitions with each phantom and institution combination. The blue shaded region corresponded to the expected tolerance of the error as determined by the manufacturer-defined physical density tolerance.

**Table 3.**
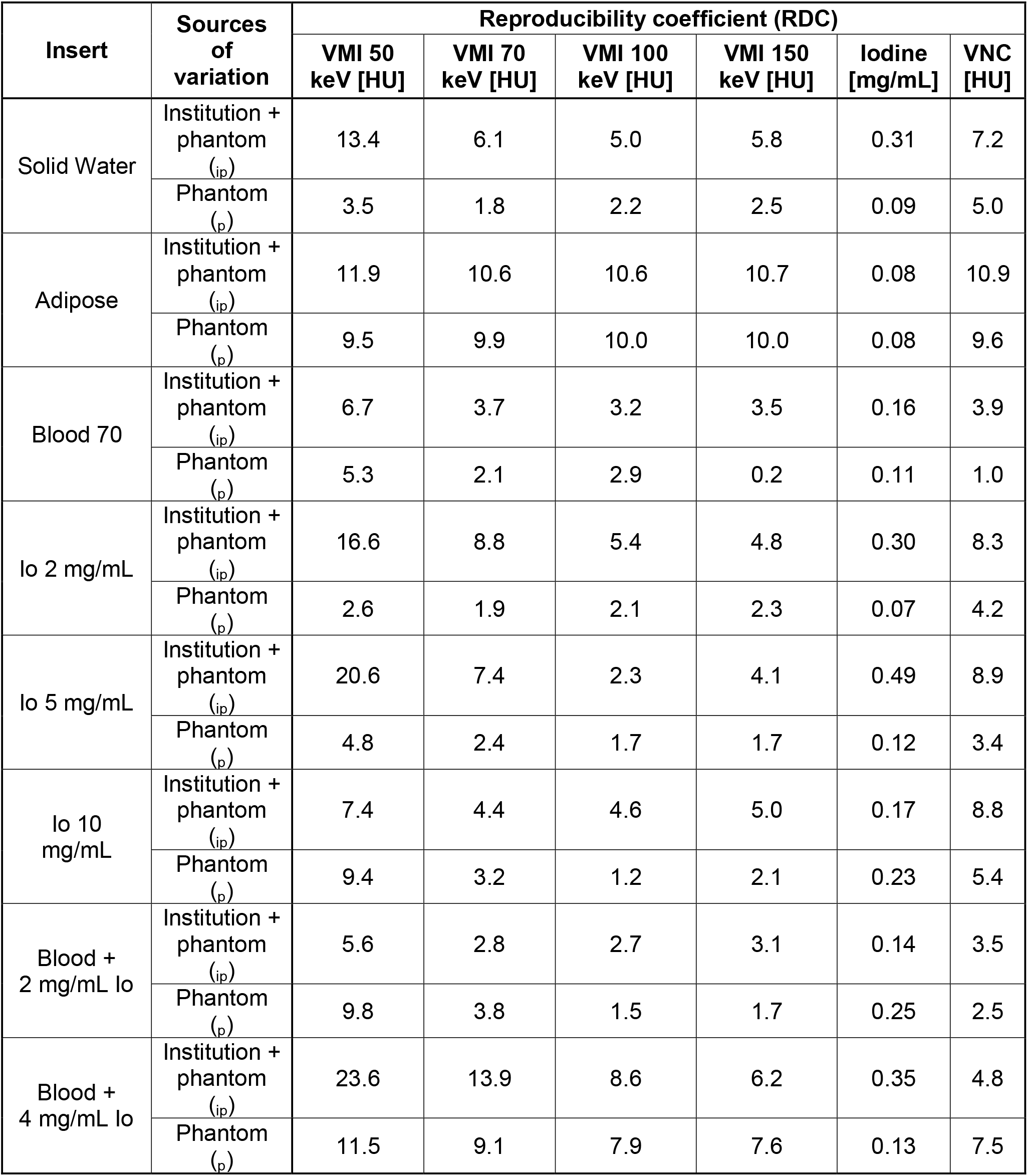
Reproducibility coefficient (RDC) for each insert and spectral results for different sources of variation.

| Insert | Sources of variation | Reproducibility coefficient (RDC) |  |  |  |  |  |
|---|---|---|---|---|---|---|---|
|  |  | VMI 50 keV [HU] | VMI 70 keV [HU] | VMI 100 keV [HU] | VMI 150 keV [HU] | Iodine [mg/mL] | VNC [HU] |
| Solid Water | Institution + phantom (<sub>ip</sub>) | 13.4 | 6.1 | 5.0 | 5.8 | 0.31 | 7.2 |
|  | Phantom (<sub>p</sub>) | 3.5 | 1.8 | 2.2 | 2.5 | 0.09 | 5.0 |
| Adipose | Institution + phantom (<sub>ip</sub>) | 11.9 | 10.6 | 10.6 | 10.7 | 0.08 | 10.9 |
|  | Phantom (<sub>p</sub>) | 9.5 | 9.9 | 10.0 | 10.0 | 0.08 | 9.6 |
| Blood 70 | Institution + phantom (<sub>ip</sub>) | 6.7 | 3.7 | 3.2 | 3.5 | 0.16 | 3.9 |
|  | Phantom (<sub>p</sub>) | 5.3 | 2.1 | 2.9 | 0.2 | 0.11 | 1.0 |
| Io 2 mg/mL | Institution + phantom (<sub>ip</sub>) | 16.6 | 8.8 | 5.4 | 4.8 | 0.30 | 8.3 |
|  | Phantom (<sub>p</sub>) | 2.6 | 1.9 | 2.1 | 2.3 | 0.07 | 4.2 |
| Io 5 mg/mL | Institution + phantom (<sub>ip</sub>) | 20.6 | 7.4 | 2.3 | 4.1 | 0.49 | 8.9 |
|  | Phantom (<sub>p</sub>) | 4.8 | 2.4 | 1.7 | 1.7 | 0.12 | 3.4 |
| Io 10 mg/mL | Institution + phantom (<sub>ip</sub>) | 7.4 | 4.4 | 4.6 | 5.0 | 0.17 | 8.8 |
|  | Phantom (<sub>p</sub>) | 9.4 | 3.2 | 1.2 | 2.1 | 0.23 | 5.4 |
| Blood + 2 mg/mL Io | Institution + phantom (<sub>ip</sub>) | 5.6 | 2.8 | 2.7 | 3.1 | 0.14 | 3.5 |
|  | Phantom (<sub>p</sub>) | 9.8 | 3.8 | 1.5 | 1.7 | 0.25 | 2.5 |
| Blood + 4 mg/mL Io | Institution + phantom (<sub>ip</sub>) | 23.6 | 13.9 | 8.6 | 6.2 | 0.35 | 4.8 |
|  | Phantom (<sub>p</sub>) | 11.5 | 9.1 | 7.9 | 7.6 | 0.13 | 7.5 |

Error for low energy VMIs displayed more variability between institutions compared to at VMI 70 keV (Figure 5). At VMI 50 keV, the σ_ip_ of the iodine 5 mg/mL insert was 7.4 HU compared to 2.6 HU at VMI 70 keV (Figure 4). σ_ip_ of the iodine 5 mg/mL insert further increased to 12.3 HU at VMI 40 keV, which was less than the corresponding σ_p_ of 2.8 HU at VMI 40 keV. Similarly, with the blood 70 insert, the σ_ip_ measured 3.9, 2.4, 1.6, and 1.3 HU compared to σ_p_ of 3.1at VMI 40, 50, 60, and 70 keV, respectively. The inter-institutional variability at low energy VMIs was pronounced, i.e. RDC_ip_ > 10 HU, in particular inserts, such as adipose, solid water, iodine 2 mg/mL, iodine 5 mg/mL, and blood + 4 mg/mL iodine. Notably, the standard deviation of the RDC_ip_ across the inserts increased from 3.8 to 6.6 HU for VMI 70 to 50 keV. On average, the RDC_ip_ for VMI 50 keV was 13.2 ± 6.7 HU in comparison to the average RDC_p_ of 7.1 ± 3.4 HU, displaying the enhanced effect of institution at lower energy VMIs (Table 3).

As VMI energy increased from 70 keV to 150 keV, variability between institutions and phantoms stabilized (Figure 5). The σ_ip_ for the iodine 5 mg/mL insert were 7.4, 2.6, 0.9, and 1.5 HU for VMI 50, 70, 100, and 150 keV, respectively (Figure 4). A similar trend was observed for all other inserts: with the solid water and blood 70 insert, the σ_ip_ decreased from 4.8 to 2.1 and 2.4 to 1.3 HU between VMI 50 and 150 keV, respectively. Across inserts, the σ_ip_ was more similar to σ_p_ at higher energy VMIs than low energy VMIs: for the iodine 5 mg/mL insert, the σ_ip_ and σ_p_ were 1.5 and 0.7 HU at VMI 150 keV and 7.4 and 1.8 HU at VMI 50 keV, respectively (Figure 4). This decrease in σ_ip_ corresponded to a decrease in the RDC_ip_ from 20.6 HU at VMI 50 keV to 4.1 HU at VMI 150 keV (Table 3). At VMI 100 and 150 keV, RDC_ip_ averaged 5.3 ± 2.9 and 5.4 ± 2.4 HU, respectively, compared to the RDC_ip_ of 7.2 ± 3.8 HU at VMI 70 keV. In comparison, the RDC_p_ across inserts was 3.7 ± 3.3 and 3.5 ± 3.4 HU for VMI 100 and 150 keV, respectively, highlighting the reduced influence of different institutions’ scanners on quantitative variability as VMI energy increased.

Iodine and VNC exhibited similar error between phantoms and between institutions and phantoms. The resulting error for the iodine 5 mg/mL inserts at each institution was 0.23 ± 0.05, 0.28 ± 0.04, 0.12 ± 0.03, and 0.54 ± 0.06 mg/mL (Figure 6A). These errors corresponded to a σ_ip_ of 0.18 mg/mL compared to the σ_p_ of 0.04 mg/mL, showing the stronger effect of institution. Across inserts, the RDC_ip_ ranged from 0.08 to 0.49 mg/mL, averaging 0.3 ± 0.1 mg/mL (Table 3). VNC error also demonstrated consistency: the error of the iodine 5 mg/mL insert at the different institutions was -2.3 ±1.1, -4.0 ± 1.2, -3.8 ± 1.3, and -9.6 ± 1.4 HU (Figure 6B). The corresponding σ_ip_ was 3.2 HU compared to the σ_p_ of 1.3 HU. For the inserts, the RDC_ip_ ranged from 3.5 to 10.9 HU.

**Figure 6.**
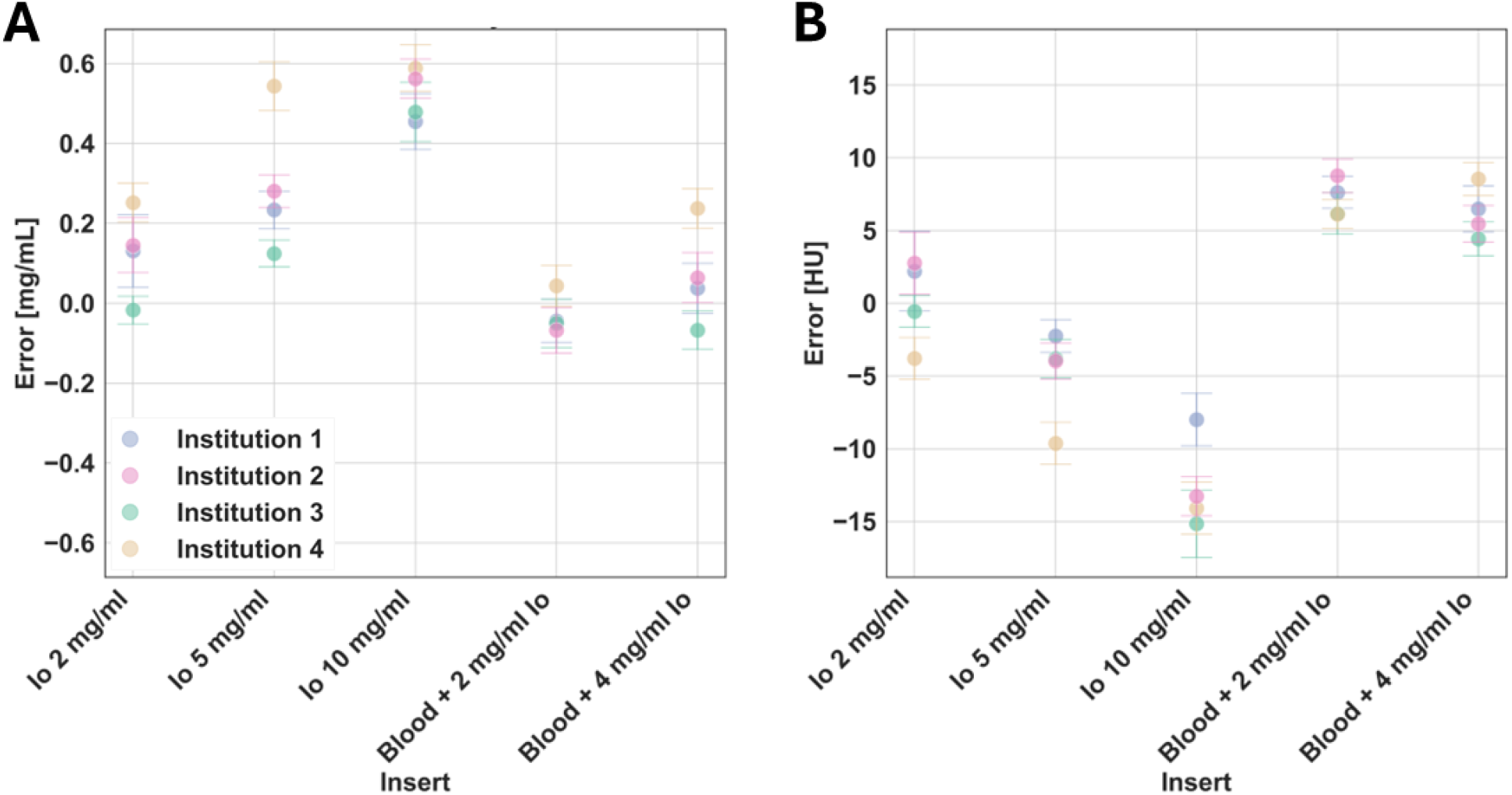
Iodine and VNC error across four institutions. Iodine error (A) showed similar values between institutions across inserts. Similarly, VNC error (B) demonstrated strong overlap in values for iodine-containing inserts. Error bars corresponded to the standard deviation across slices and scan repetitions for a given insert, phantom, and institution combination.

## Discussion

PCCT demonstrated quantitative consistency for all spectral results across institutions using the same scanner model, phantom model, and protocol with average RDC_ip_ ranging from 5.4 to 13.2 HU for VMIs and VNC and RDC_ip_ of 0.3 mg/mL for iodine maps. As VMI energy increased, the variability between institutions and phantom copies decreased before stabilizing to an average RDC_ip_ of 5.4 HU. Variability from institutions particularly dominated over phantom variability in low energy VMIs as demonstrated by an average increase of 6.2 HU from RDC_p_ to RDC_ip_ for VMI 50 keV. Overall, PCCT-derived spectral results exhibited intra-institutional repeatability and inter-institutional reproducibility that will facilitate the development of reproducible and clinically useful biomarkers.

Unlike the few studies that examined intra-scanner model variability at a single institution^40,41^, this study determined the inter-institutional variability. Inter-institutional variability incorporates not only intra-scanner model variability but also potential variability stemming from differences in ambient operating conditions that may impact the sensitive photon-counting detector. In a single center study with the same PCCT model utilized in this study, VMI 70 keV quantification in a lung phantom was within 3 HU between two scanners with matched acquisition and reconstruction parameters^41^. Comparatively, our study observed a larger range of values across institutions with a corresponding RDC_ip_ of 7.2 ± 3.8 HU at VMI 70 keV. The reduced reproducibility in our study can likely be attributed to both phantom variability and the inclusion of PCCTs across the United States instead of at a single institution. In other spectral CT systems, intra-scanner model variability has only been evaluated in a kVp switching CT^40^. Across 11 copies of the same scanner model, the intra-scanner model standard deviation was 0.16 and 0.18 mg/mL for the iodine 2 mg/mL and iodine 5 mg/mL insert, respectively^40^. This variability matched our σ_ip_ of 0.11 and 0.18 mg/mL for the iodine 2 mg/mL and iodine 5 mg/mL inserts, respectively. However, our study also includes phantom variability in the σ_ip_, suggesting that intra-scanner model variability with PCCT may be less compared to other spectral CT systems. For other CT systems, only inter-scanner model and intra-scanner comparisons have been performed^17,42–47^.

The inter-institutional reproducibility fundamentally bolsters prior phantom and clinical studies to support continued clinical translation of PCCT. While this study only characterized inter-institutional reproducibility of a single protocol, the variation between four institutions is well defined across the spectral results. As a result, this captured variation may translate to other previously published studies that have highlighted the potential benefit of quantification with PCCT but only on a single system. The known variation across spectral results provides context for inter-institutional implementation and boosts confidence in quantification to encourage further adoption of PCCT clinical applications.

In conjunction with growing confidence in the quantitative stability across protocol parameters, the inter-institutional reproducibility of PCCT promotes the development and clinical adoption of PCCT-derived biomarkers. In order to achieve widespread clinical adoption, these biomarkers must be generalizable between institutions, between protocol parameters, and over time to isolate biological disease from non-biological sources of variation^48^. Despite investigations of biomarker variability^49,50^, there are no established benchmarks or standards that affirm the quantitative reproducibility required for biomarkers. As a result, characterization of the inter-institutional reproducibility can provide guidance in the development of imaging biomarkers and provides variability information required for reliable biomarker quantification. This direct incorporation of variability in biomarker design ensures accurate and consistent diagnostic interpretation across patients and longitudinally detection of nuanced changes in disease state. This approach varies from others commonly used methods in biomarker evaluation^51–54^, including both statistical^55,56^ and deep learning harmonization^57–59^, which attempt to correct for sources of variation but require copious amounts of data to establish a model. While this study only evaluated a single protocol at a single time point, PCCT has demonstrated quantitative stability, i.e. VMI 70 keV values within 10 HU, across key parameters, including patient size, radiation dose level, reconstruction kernel, and denoising^25–27,30,31,60–63^. In addition, PCCT has displayed quantitative stability over time with variation in iodine density ranging from 0.02 to 0.07 mg/mL and variation in VMI 70 keV relative error averaging 0.11% across two years^34^. As a result, our study provides a baseline understanding of inter-institutional reproducibility in addition to variability from other parameters that can be included in biomarker development and evaluation. To date, only a single study has demonstrated both intra-scanner model and protocol parameter variability simultaneously, showing VMI 70 keV quantification falling within 8 HU with different dose levels, reconstruction kernels, and denoising levels^34^.

This study has some limitations. First, the same copy of the phantom was not scanned at each of the four institutions. As a result, the analysis combines both phantom- and scanner-related variation. To account for phantom-related variation, slight differences in insert composition and physical density were utilized to calculate the expected value for each institution’s phantom. In addition, inserts from each institution’s phantom were scanned at a single institution to isolate this effect from inter-institutional effects. Second, only four institutions participated in the study. However, the four institutions were in varying climates across the United States. Future studies will include additional institutions to further refine the inter-institutional reproducibility. In addition, the study can be extended over time to further track the quantitative reproducibility over time as well as examine the effect of climate, room temperature, and humidity across systems. Of note, at the time of the study, the four institutions did not cite any specific variations in room temperature and humidity. Third, only one set of protocol parameters were utilized in order to provide a baseline inter-institutional reproducibility. Future work will assess reproducibility across key sources of variation, such as patient size and tube voltage. Last but not least, this study performed scans on a singular clinical PCCT model. At the time of the study, only one scanner model has been approved for clinical use. Future studies will incorporate other PCCT systems to fully delineate reproducibility across different scanner models.

## Conclusion

PCCT-derived spectral results were quantitatively repeatable and reproducible across four institutions with RDC_ip_ ranging from 5.4 ± 2.4 to 13.2 ± 6.7 HU for VMIs and VNC and a RDC_ip_ of 0.3 ± 0.1 mg/mL for iodine maps. The reproducibility across institutions boosts confidence in previously established benefits of PCCT and supports its continued clinical translation and adoption. In addition, the consistent quantification enables reproducible diagnostic interpretation with a given protocol. Characterization of inter-institutional variability provides critical information for the development of generalizable biomarkers with standardized acquisition protocols. This work demonstrates that the quantitative reproducibility of PCCT encourages the continued development and clinical adoption of biomarkers for a wide range of applications.

## Data Availability

All data produced in the present study are available upon reasonable request to the authors.

## Author contributions

LL, PN, and MM contributed to the conception of the study. LL, MR, PN, LR, XD, KR, DZ, GK, and MM designed the study parameters. LL, LR, KR, and MM acquired image data. LL and GK performed analysis of the phantoms. LL, GK, and MM drafted the manuscript. All authors revised and reviewed the manuscript before submission.

## Abbreviations

VMI: virtual monoenergetic images
VNC: virtual non-contrast
PCCT: photon-counting CT
HU: Hounsfield units
ROI: region of interest
RC: repeatability coefficient
RDC: reproducibility coefficient

